# A prospective cohort study to assess if alcohol intake measured by routine pregnancy self-report predicts developmental concerns uncovered by routine health visitor screening of children at 30 months of age

**DOI:** 10.1101/2025.01.08.25320200

**Authors:** David Tappin, Daniel Mackay, Lucy Reynolds, Niamh Fitzgerald

## Abstract

**Background:** Stigmatized behaviours are often underreported, especially in pregnancy, making them challenging to address. The Alcohol and Child Development Study (ACDS) seeks to inform prevention of foetal alcohol harm, linking self-report as well as a maternal blood alcohol biomarker with child developmental outcomes.

**Methods:** Maternity records from all pregnant women in the study city who presented for maternity care during the 12-month period June 2017 – June 2018 were transferred to the safe data facility creating the baseline cohort. Health Visitor routinely recorded child developmental screening data collected when the offspring were 30 months of age were transferred and linked to the cohort. Anonymous analysis was performed to assess associations between self-reported alcohol intake collected at maternity presentation with child development concerns and looked after by the local authority status of offspring at 30 months of age. Chi2 tests of association were used.

**Results:** 14919 maternity records were transferred and 10876 could be linked to developmental screening data collected at 30 months of age. As expected self-reported current alcohol use after conception measured at presentation to maternity care was associated with offspring being looked after by the local authority (chi^2^=7.85, p=0.005) at age 30 months. 6.6% of children in care were attributable to self-report of current alcohol use during pregnancy. Developmental outcomes were generally inversely associated with self-report of current alcohol use notably concern regarding Social Development (chi^2^=4.08, p=0.043) and Speech, Language or Communication Development (chi^2^=4.37, p=0.037).

**Conclusions:** Explanation for this unexpected inverse relationship may relate to known miss-representation of alcohol intake from maternal self-report (the risk) or inaccurate assessment of offspring development at 30 months (the outcome). However self-report of current intake at maternity presentation prompts an alcohol intervention which if effective might explain the anomalous findings.

## Introduction

Foetal Alcohol Syndrome (FAS) is recognized in the developed world as the leading preventable cause of disorders of intellectual development (1). According to the literature, the overall combined rate of FAS and Fetal Alcohol Spectrum Disorders (FASD) is estimated to be about 7-18 /1,000 births in various populations (2). Both the diagnosis of FAS/FASD soon after birth (3) and the direct measurement of alcohol in the mother are difficult (4,5), posing challenges for the evaluation of interventions aimed at reducing dangerous antenatal alcohol consumption. Most cases of developmental impairment that are probably caused by alcohol use during pregnancy are never known to health carers to be related to alcohol use.

Currently all pregnant women are asked about alcohol use by their midwife at maternity presentation in early pregnancy (6). Support offered ranges from a brief intervention (7) to referral for additional support from a specialist nursing team depending on level of alcohol use. However, a recent study showed that self-report only identified 38% of women who drank significant alcohol during pregnancy (8). False negative information is given for a variety of reasons: social stigma, inaccurate recollection and problems estimating alcohol content or volume consumed with consequent under-reporting (9). Self-report is also influenced by a desire to provide socially acceptable information (10) or for fear of censure and intervention by social services (11). Most other treatable conditions that might be hidden or unknown like smoking and hepatitis B are tested for using a breath test and blood test respectively, so that all can be offered effective therapy or treatment. There is currently no recognised test to augment self-report of alcohol use during pregnancy. This means that specific treatment for the mother, her child and prevention of damage for future children is often not provided.

The Alcohol and Child Development Study (ACDS) involves a cohort of pregnant women in a UK city and aims to inform the development of improved methods of identification of women at risk of having a child affected by foetal alcohol exposure. The study aims to follow anonymously the children of a full annual cohort of women identified at their first maternity visit. Examination for a relationship will be made between self-reported alcohol consumption in pregnancy and/or a positive maternal blood test for an alcohol biomarker and later childhood developmental impairment. Developmental impairments have been taken from routine health visitor developmental screening at 30 months of age and will also be derived from routine clinical assessment of any of the cohort of children referred for suspected developmental problems requiring specialist assessment and treatment pre-school.

This paper reports self-report of alcohol use before and during early pregnancy, and referral for support. Associations with developmental concerns discovered during routine developmental screening by health visitors at 30 months postnatal age are reported.

## Methods

The Alcohol and Child Development Study (ACDS) involves a cohort of pregnant women in a UK city and aims to inform the development of improved methods of identification of women at risk of having a child affected by foetal alcohol exposure. The study anonymously follows the children of a cohort of women identified in pregnancy, to examine whether or not there is a relationship between either positive maternal blood tests for alcohol biomarkers and/or self-reported alcohol consumption in pregnancy, and later childhood developmental impairment.

### Design

This paper compares routinely collected data related to alcohol use before and during pregnancy from women attending ‘booking’ appointments for maternity care in the study city, with later routine screening child development information collected at 30 months of age by health visitors.

### Setting

The setting for the ACDS was midwife-led first maternity clinic appointments at 13/14 weeks of pregnancy in a medium sized UK city between June 2017 and June 2018 (‘the study period’). These appointments are to start the midwife-led pregnancy support programme and include a formal pregnancy test, breath test for smoking as well as routine blood tests for infection with hepatitis B, syphilis and HIV for which current treatment improves birth outcomes. Women are also asked about alcohol use. If significant alcohol consumption is reported, women are provided with a brief intervention or referral to specialist services to support alcohol reduction and elimination during pregnancy.

### Participants and information provided

All pregnant women reporting to the NHS in the study city during the study period received a study information sheet (PIS) along with the letter informing them of their booking appointment date and time and all women who attended during the analysis period are included in the analysis for this paper as outlined above. In line with the ethical approval conditions, the PIS informed women that the NHS wished “to collect an extra 2ml (one third of a teaspoon) of blood from all pregnant women”, and that this extra blood would be “used anonymously for research … which may help us to learn more about the best ways to support the health of women and babies and improve our service.” Women were informed that they could opt out of the study by letting their midwife know that they did not want the sample taken. A contact name and number was provided on the PIS for any queries.

Similar generic information was provided to all midwives by a senior research midwife unconnected to the study team, and they were advised on when and how to take the additional sample. Midwives were asked not to actively consent the women before taking the extra sample, but to take the sample ‘routinely’ unless the woman (from reading the PIS sent with the booking appointment notification) voiced a wish to opt out. The potential testing of the additional blood sample for an alcohol biomarker was not disclosed to women or frontline midwives.

The blood samples were stored in a sample repository with a study identifier linked to routine maternity booking data held by a secure NHS data service such that neither the researchers nor clinicians could identify who had a sample taken. The study aimed to assay these samples for a marker used by the Driver and Vehicle Licensing Agency (https://www.drinkdriving.org/cdt-alcohol-test.php) to assess current heavy alcohol intake– Carbohydrate Deficient Transferrin (12).

### Quantitative variables and data sources

Detailed self-report of alcohol use was collected at the first maternity visit via a locally developed computerised data system until 31st October 2017. A change in computer system from November 2017 meant that less detailed self-reported data was recorded, through to the end of sample collection in June 2018, regarding alcohol use during pregnancy. With the exception of birth weight of the baby, variables were gathered from routinely collected maternity service data recorded directly by midwives onto a computerised database. Only those variables which were deemed not to risk the anonymity of participants were shared with researchers. Shared data included the following:

- Area-based material deprivation, (13) calculated from patient postcode using government statistics.
- Age
- Height and weight at the time of booking, used to calculate Body Mass Index (BMI) using the formula Weight (in kilos)/Height2 (in metres).
- Number of previous pregnancies, previous spontaneous abortions, previous therapeutic abortions.
- Smoking history, self-reported smoking while pregnant, carbon monoxide breath test level
- Estimated gestation in weeks calculated from recall of last menstrual period, and
- Self-reported alcohol use. This was recorded in 16 domains, of which just four of these contained sufficient data for the safe data facility to be sure that anonymity would be retained. How much did you drink daily before pregnancy? (converted into alcohol units); Brief Intervention was required Yes/No?; Referral to (alcohol) intervention nurse Yes/No?; How much do you drink each week now? (converted into alcohol units).
- The birth weight of the baby was obtained through record linkage with a national database.

All of this data was transferred to and held by an NHS safe data facility to enable future linkage with child health records without compromising the anonymity of the participants or their children.

### Outcome data

The health service aims for all children born in Scotland to have a developmental assessment by Health Visitors gathering data from parents/carers at 27 to 30 months postnatal age, as part of a national screening programme (14,15). At this contact questions included assessment of: Looked After Child status; Carer’s Smoking status; Child Exposed to ETS (Environmental Tobacco Smoke); and developmental achievements: Social Development; Emotional Development; Speech, Language and Communication Development; Gross Motor Development; Fine Motor Development; Vision; Hearing. Parental responses were recorded as: New concern; Previous concern; No concern; No meaningful result. Health Visitors complete an assessment template on a computer-based data collection system (EMIS) and then this data is extracted to the national data set (15). Data held by the health board for children born between 1^st^ August 2017 and 28^th^ February 2019 were provided to the safe data facility to include pregnancies where women booked for maternity care between 12th June 2017 and 30th June 2018. Most data was collected at 30 months postnatal age during 2020 when the percentage of children from whom collection was complete are shown below.

### Bias

The unusual consent procedures in this study described above – passive consent for generic research using a routine extra blood sample - were designed to minimise sources of bias in the cohort. Bias and ethical issues have been discussed previously (16).

### Study size

The whole cohort from 12th June 2017 to 30th June 2018 was chosen to include all festivals and holiday periods during a full calendar year when alcohol intake may increase.

### Statistical methods

Statistical analyses were performed with Stata 12.10 [17]. Descriptive statistics (percentages, medians and interquartile ranges) were derived for all variables. Logistic regression was used to determine whether alcohol intake predicted non-delivery of a blood specimen after controlling for age category and deprivation quintile.

### Ethics

Ethics approval was granted by the local NHS Research Ethics Committee (10/06/2014 - approval number provided, but not published to protect the identity of the study site). This has been previously discussed in detail (16).

Permission for outcome data, from the 27-30 month Health Visitor developmental assessment held by the health board, to be passed to the safe data facility was given by the Director of Public Health. Ethics approval for linking this data with the maternity booking data was provided by the ethics committee within the safe data facility (03/11/2021).

The sponsor for study was the local NHS management authority.

## Results

Self-reported alcohol use data for the cohort of pregnant women who booked for maternity care in the city between 17^th^ June 2017 and the 30^th^ June 2018 is shown in table 2. How much did you drink daily before pregnancy? (converted into alcohol units); Brief Intervention was required Yes/No?; Referral to (alcohol) intervention nurse Yes/No?; How much do you drink each week now? (converted into alcohol units). At maternity booking if a pregnant woman indicates that she is currently drinking 15 units per week or more, a referral to the Special Needs In Pregnancy service is required. For 1-14 units an Alcohol Brief Intervention provided by the attending midwife is required.

**Table 1.** Percentage of children where 27-30 review data were collected by calendar month for 2020.

| **Uptake 2020** | **Jan** | **Feb** | **March** | **April** | **May** | **June** | **July** | **Aug** | **Sept** | **Oct** | **Nov** | **Dec** |
| --- | --- | --- | --- | --- | --- | --- | --- | --- | --- | --- | --- | --- |
| **% 30 Month Completion** | **94%** | **93%** | **91%** | **85%** | **72%** | **57%** | **65%** | **76%** | **84%** | **88%** | **90%** | **92%** |

**Table 2.**

| **Alcohol units per day before conception** |  |
| --- | --- |
| Units | Freq |
| 0 | 10724 |
| 1-4 | 1247 |
| 5-14 | 90 |
| >=15 | 21 |
| Not asked or not known | 1348 |
| Total | 13430 |
| **Alcohol units per week now/since conception** |  |
| Units | Freq |
| 0 | 12099 |
| 1-4 | 402 |
| 5-14 | 124 |
| >=15 | 22 |
| Not asked or not known | 783 |
| Total | 13430 |
| **Brief intervention given** |  |
| Yes | 31 |
| No | 13399 |
| Total | 13430 |
| **Referred to alcohol intervention nurse/team** |  |
| Yes | 246 |
| No | 13184 |
| Total | 13430 |

We chose to use the alcohol variable: Weekly alcohol since conception Yes/No (if data MISSING then daily alcohol pre-conception Yes/No was used as surrogate)’.

Frequencies of observations for 27-30 month developmental assessment are shown in supplementary table 1 for 10,876 children where data was recorded. There were good levels of completion (92-99%) for nearly all variables examined which were: Looked After Child status; Carer’s Smoking status; Child Exposed to Environmental Tobacco Smoke; Social Development; Emotional Development; Speech, Language and Communication Development; Gross Motor Development; Fine Motor Development; Vision; Hearing. Ages and Stages Questionnaire (ASQ) was only completed for 428(4%) of children and was not examined further.

Cross-tabulation of these outcomes with ‘Alcohol use now or since conception’ showed that Social Development (chi2 = 4.08, p=0.04) Table 3a had a greater proportion of concerns among women where <u>no</u> alcohol use was reported during pregnancy, as did concerns regarding Speech, Language, Communication Development (chi2 = 4.37, p=0.04) Table 3b.

**Table 3.**
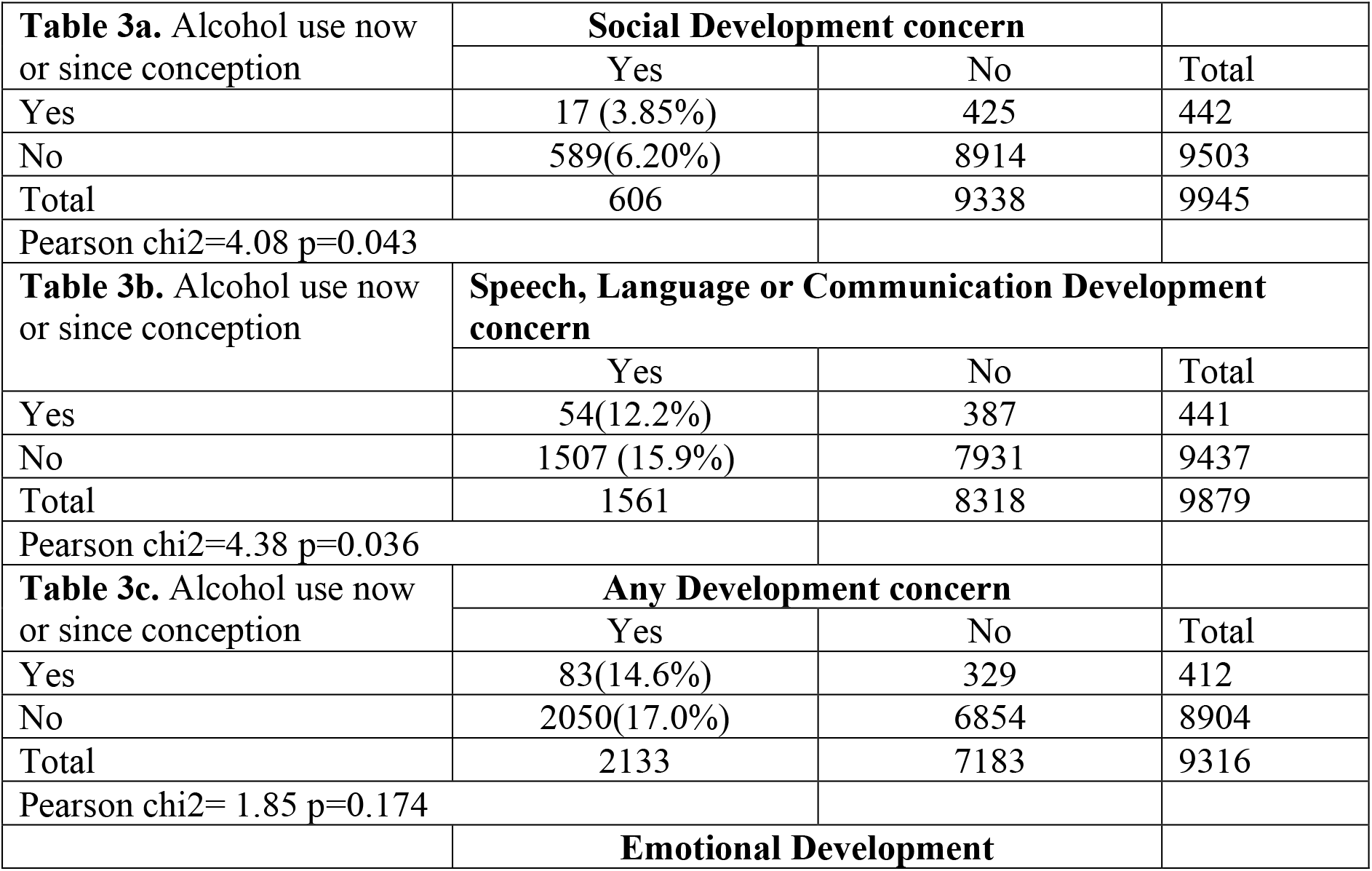
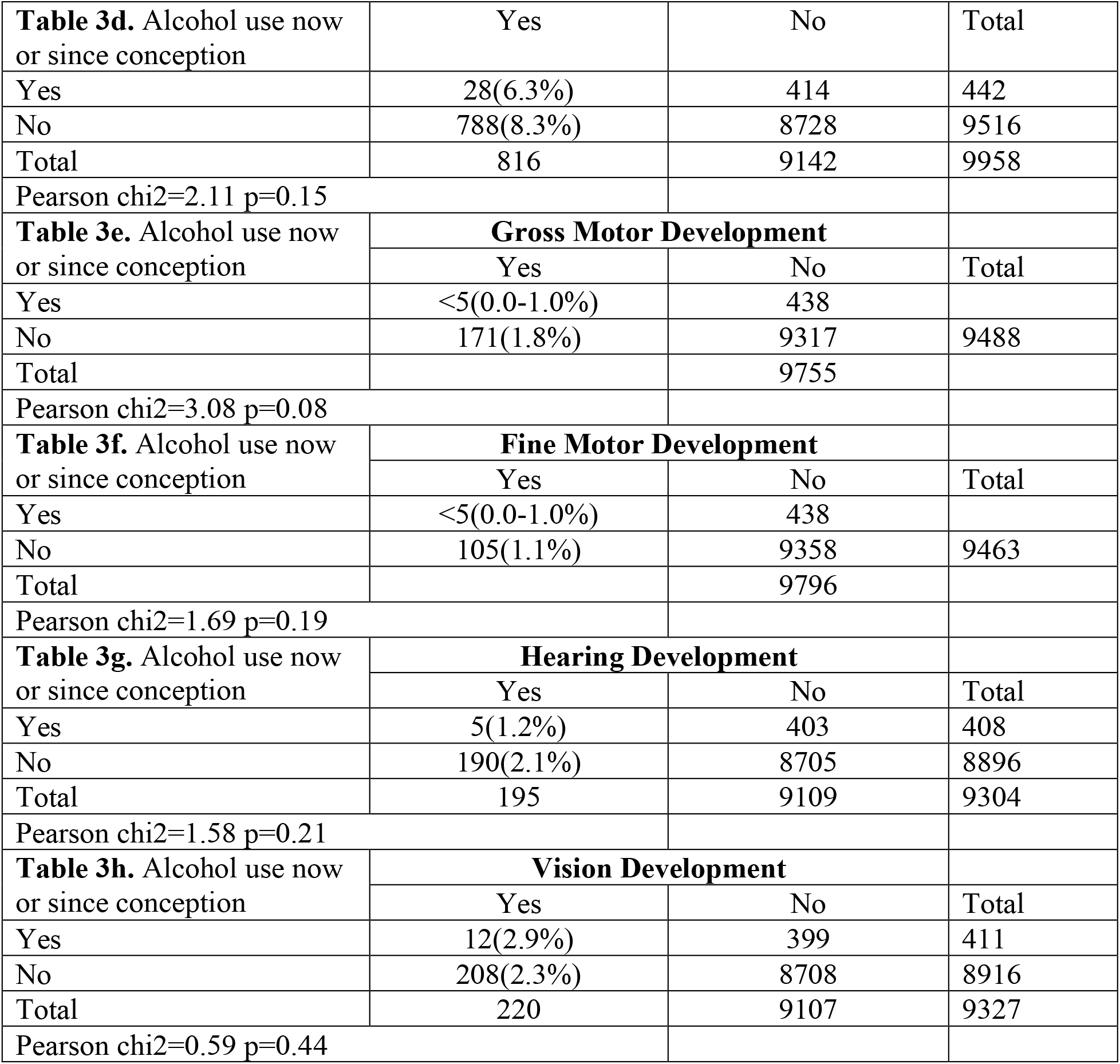

Any Developmental Concern showed no-significant association (chi2 = 2.19, p=0.14) table 3c, as did Emotional Development (chi2 =2.11, p=0.147) table 3d, Gross Motor Development (chi2 = 3.08, p=0.08) table 3e, Fine Motor Development (chi2 = 1.69, p=0.19) table 3f, Hearing Development (chi2 = 1.58, p=0.21) table 3g but all these developmental outcomes were in the direction of more concerns within the self-report <u>no</u> alcohol since conception group. Vision Development (chi2 = 0.59, p=0.44) table 3h was not statistically significant but was in the direction of more concerns for those self-reporting alcohol use now or since conception.

The only data item with an ‘expected’ (18) statistically significant association with reported alcohol use now or since conception was Looked After (by the local authority) shown in table 4.

**Table 4.**

| Alcohol units per week now/since conception | Looked After (by the local authority) |  | Total |
| --- | --- | --- | --- |
|  | Yes | No |  |
| Yes | 9 (2.01%) | 439 | 448 |
| No | 75 (0.78%) | 9563 | 9638 |
| Total | 84 | 10002 | 10086 |

Pearson $\chi^2=7.85$ $p=0.005$
Attributable Risk = $a/(a+b) - c/(c+d) = 9/448 - 75/9637 = 0.0123$
PAR = $AR \times P_e = 0.0123 \times 448/10085$
PAR% = $PAR/Incidence \text{ of Looked after} \times 100 = (0.0123 \times 448/10085)/(84/10085) = 6.6\%$

Incidence of being ‘Looked After’ in whole population = 84/10085 Attributable Risk = a/(a+b) – c/(c+d) = 9/448 – 75/9637 = 0.0123 PAR= AR × P_e_ = 0.0123 × 448/10085

PAR% = PAR/Incidence of Looked after × 100 = (0.0123 × 448/10085)/(84/10085) = 6.6%

This result suggests that for about 7% of children looked after by the local authority this is attributable to self-report of current alcohol use at presentation for maternity care.

## Discussion

This study indicates that ‘self-reported’ alcohol use during pregnancy disclosed at maternity booking, usually in the second trimester, is inversely associated with concerns about some developmental outcomes collected by health visitors from parents at 30 months postnatal age. This is a surprising outcome as it contradicts most other evidence that maternal alcohol use during pregnancy is detrimental to child development outcomes (19,20).

### Possible explanations

There are a number of possible reasons for these interesting findings that can be split into 3 categories:

1. The measurement of ‘risk’: Maternal alcohol use at maternity presentation by ‘self-report’ is known to miss-report many women who drink alcohol during pregnancy (8) because of perceived fear of judgement by health service staff and even social services intervention to protect the child once born. This is backed up by the only outcome not relying on maternal self-report, ‘Looked-after’ status, which shows an expected positive relationship with alcohol use during pregnancy.
2. Measurement of ‘outcome’: Health Visitor led developmental assessment at 30 months postnatal age, although structured, relies on maternal self-report which may be inaccurate at predicting poor outcomes. ‘Effectiveness of screening’ assessment for this type of child development screening pre-school is yet to be undertaken. Up to 10% of children do not undertake this developmental screening programme (21) and the most in need are likely to be over-represented in the ‘unscreened’.
3. There is an inverse association between some developmental concerns at 30 months and self-reported current alcohol use documented at maternity booking because pregnant women who self-reported alcohol use received an effective intervention (either a brief intervention from midwifery staff for any reported current alcohol intake or more extensive support from specialist nurses if current reported alcohol intake was high). Drinking alcohol may have stopped for the rest of these pregnancies and developmental concerns in babies were averted. This possibility is supported by those <u>not</u> reporting current alcohol use having more developmental ‘concerns’ particularly related to ‘social development’ and ‘speech language or communication development’ which are areas often reported with alcohol related damage (22). Those who self-report as <u>no</u> current alcohol use will include a proportion of women who are ‘miss-reporting’ and are in fact current alcohol users at maternity booking. These women will therefore not receive an ‘effective’ intervention to stop drinking alcohol at maternity presentation, whereas those who self-report current alcohol use received an ‘effective’ intervention and therefore stopped drinking alcohol.

In order to resolve possibility 1., a better method of documenting excessive alcohol use preferably before pregnancy, but at least early in pregnancy is required. This study has collected extra blood samples to test anonymously for an alcohol metabolite at maternity booking. Once these samples are assayed, the results can be linked in the safe data facility to self-report of current alcohol use and developmental outcome measures.

To resolve possibility 2., a better outcome measure would be referral (often from health visitors or pre-school nursery placement) for assessment by community-based child assessment services including: speech and language therapists, specialist nurses and paediatricians in child development teams, physiotherapists, occupational therapists, hearing and vision assessment, psychologists, psychiatrists or the health team involved in fostering and adoption services. Observation over time in a nursery setting which graduates children towards the setting required for successful school placement misses few children who require assessment and support. Referrals are made in as timely a way as possible so that all children receive help and assessment prior to primary school placement. Documentation of referrals and subsequent assessments by community services are held on EMIS databases. Linkage of HV screening at 30 months with outcomes up to school age within this cohort will also assess the sensitivity and specificity of the 27-30 month developmental screening undertaken by Health Visitors.

For the third possibility, if the ‘Gold Standard’ developmental outcome discussed’ above is in general agreement with the 30-month developmental screening by health visitors, then it becomes clearer that obstetric staff are utilising an effective intervention strategy to reduce self-reported alcohol use at maternity booking. It therefore becomes more pressing to identify all pregnancy alcohol use at maternity presentation. By testing the stored serum samples for Carbohydrate Deficient Transferrin (which has low sensitivity but high specificity), the size of under-reporting of current alcohol use at maternity booking in this cohort of pregnant women can be estimated.

### Further research using this cohort

This cohort study could add important information regarding the missed opportunity to intervene by assaying the stored samples for an alcohol metabolite, Carbohydrate Deficient Transferrin (CDT) which has high specificity and low sensitivity. Linkage of CDT results to self-report of current alcohol use at maternity booking will show the proportion self-report positive results that are also CDT positive. Then the number of false self-report negative current alcohol use responses can be estimated from the remaining CDT positive results allowing the proportion of missed opportunities to intervene to be calculated.

A possible ‘gold standard’ development ‘outcome’ for this and other similar cohorts can be examined by transferring health board pre-school developmental assessment and support data held on EMIS databases to the safe data facility for linkage and anonymous analysis.

## Data Availability

All data produced in the present work are contained in the manuscript

## References

1. McCarthy R, Mukherjee RAS, Fleming KM, Green J, Clayton-Smith J, Price AD, Allely CS, Cook P A. Prevalence of fetal alcohol spectrum disorder in Greater Manchester, UK: An active case ascertainment study. Alcoholism: Clinical and Experimental Research 2021;45:2271–81. 10.1111/acer.14705

2. Lange S, Probst C, Gmel G, Rehm J, Burd L, Popova S. Global prevalence of fetal alcohol Spectrum disorder among children and youth. JAMA Pediatr. 2017;171(10):948–56.

3. Chasnoff IJ, Wells AM, King L. Misdiagnosis and missed diagnoses in foster and adopted children with prenatal alcohol exposure. Pediatrics. 2015;135(2):264–70.

4. McQuire C, Paranjothy S, Hurt L, Mann M, Farewell D, Kemp A. Objective measures of prenatal alcohol exposure: a systematic review. Pediatrics. 2016. 10.1542/peds.2016-0517.

5. Howlett H, Abernethy S, Brown NW, Rankin J, Gray WK. How strong is the evidence for using blood biomarkers alone to screen for alcohol consumption during pregnancy? A systematic review. Eur J Obstet Gynecol Reprod Biol. 2017. 10.1016/j.ejogrb.2017.04.005.

6. WHO. Guidelines for the identification and management of substance use and substance use disorders in pregnancy. Geneva: World Health Organization; 2014. https://apps.who.int/iris/bitstream/handle/10665/107130/9786162715266-tha.pdf?sequence=5&isAllowed=y (Accessed ?)

7. Popova S, Dozet D, Pandya E, Sanches M, Brower K, Segura L, Ondersma SJ. Effectiveness of brief alcohol interventions for pregnant women: a systematic literature review and meta-analysis. BMC Pregnancy Childbirth 23, 61 (2023). 10.1186/s12884-023-05344-8.

8. Corrales-Gutierrez I, Gomez-Baya D, Leon-Larios F, Medero-Canela R, Marchei E, Mendoza-Berjano R, García-Algar Ó. Alcohol Consumption Assessed by a Biomarker and Self-Reported Drinking in a Sample of Pregnant Women in the South of Europe: A Comparative Study. Toxics 2023;11(11):930. 10.3390/toxics11110930

9. Lange S, Shield K, Koren G, Rehm J, Popova S. A comparison of the prenatal alcohol exposure obtained via maternal self-reports versus meconium testing: a systematic literature review and meta-analysis. BMC Preg Childbirth 2014, 14:127. http://www.biomedcentral.com/1471-2393/14/127

10. Jones SC, Telenta J, Shorten A, Johnson K. Midwives and pregnant women talk about alcohol: What advice do we give and what do they receive? Midwifery 2011;27(4):489–96.

11. Stone R. Pregnant women and substance use: fear, stigma, and barriers to care. Health Justice 2015;12(3):2. doi: 10.1186/s40352-015-001.5-5.

12. Wolf K, Gross S, Marshall EJ, Walsham N, Keaney K, Robson N, Sherwood RA. Carbohydrate defcient transferrin (CDT) as a biomarker to assess drinking in high-risk drink drivers. Adv Clin Toxicol. 2019. 10.23880/act-16000160

13. Kearns A, Gibb K, Mackay D. Area deprivation in Scotland: a new assessment. Urban Stud. 2000;37(9):1535–59.

14. https://www.gov.scot/publications/scottish-child-health-programme-guidance-27-30-month-child-health-review/

15. https://publichealthscotland.scot/media/12876/2022-04-26-early-child-development-publication-report.pdf

16. Tappin D, Mackay D, Reynolds L, Fitzgerald N. Minimizing sample bias due to stigmatized behaviours: the representativeness of participants in a cohort study of alcohol in pregnancy. BMC Med Res Methodol 2022;22:138. 10.1186/s12874-022-01629-2

17. StataCorp. Stata statistical software: release 12. College Station, TX: Stata-Corp LP; 2011.

18. Tenenbaum A, Mandel A, Dor T, Sapir A, Sapir-Bodnaro O, Hertz P, Wexler ID. Fetal alcohol spectrum disorder among pre-adopted and foster children. BMC Pediatr 2020; 20: 275. 10.1186/s12887-020-02164-z

19. Scott S, Sher J. Effect of alcohol during pregnancy: a public health issue. Lancet 2023; 8(1):E4–5.

20. Broccia M, Munch A, Hansen BM, Sørensen KK, Larsen T, Strandberg-Larsen K, Gerds TA, Torp-Pedersen C, Kesmodel US. Heavy prenatal alcohol exposure and overall morbidities: a Danish nationwide cohort study from 1996 to 2018. Lancet 2023;8(1):E36–46.

21. https://publichealthscotland.scot/publications/child-health-pre-school-review-coverage/child-health-pre-school-review-coverage-2022-to-2023/

22. Subramoney S, Eastman E, Adnams C, Stein DJ, Donald KA. The Early Developmental Outcomes of Prenatal Alcohol Exposure: A Review. Front Neurol 2018; 9 doi= 10.3389/fneur.2018.01108.

