## Supplemental Table 1 for "A prospective cohort study to assess if alcohol intake measured by routine pregnancy self-report predicts developmental concerns uncovered by routine health visitor screening of children at 30 months of age"

**Supplementary Table 1: Frequency of outcomes from 27-30 month developmental assessment by health visitors**

| **Looked after by the local authority** | **Freq** | **Percent** |
| --- | --- | --- |
| Not Looked After | 10764 | 98.97 |
| Looked after | 95 | 0.87 |
| Not known | 17 | 0.16 |
| Total | 10876 | 100.00 |
| **Carer’s smoking status** | **Freq** | **Percent** |
| Non-smoker | 9515 | 87.49 |
| Smoker | 1352 | 12.43 |
| Not known | 9 | 0.08 |
| Total | 10876 | 100.00 |
| **Child exposed to exhaled tobacco smoke (ETS)** | **Freq** | **Percent** |
| No ETS exposure | 10157 | 93.39 |
| ETS exposure | 709 | 6.52 |
| Not known | 10 | 0.09 |
| Total | 10876 | 100.00 |
| **Social Development** | **Freq** | **Percent** |
| New concern | 415 | 3.82 |
| Previous concern | 239 | 2.20 |
| No concern | 10059 | 92.49 |
| No meaningful result | 163 | 1.50 |
| Total | 10876 | 100.00 |
| **Emotional Development** | **Freq** | **Percent** |
| New concern | 625 | 5.75 |
| Previous concern | 251 | 2.31 |
| No concern | 9848 | 90.55 |
| No meaningful result | 152 | 1.40 |
| Total | 10876 | 100.00 |
| **Speech, Language and Communication Development** | **Freq** | **Percent** |
| New concern | 1225 | 11.26 |
| Previous concern | 447 | 4.11 |
| No concern | 8964 | 82.42 |
| No meaningful result | 240 | 2.21 |
| Total | 10876 | 100.00 |
| **Gross Motor Development** | **Freq** | **Percent** |
| New concern | 65 | 0.60 |
| Previous concern | 120 | 1.70 |
| No concern | 10508 | 98.32 |
| No meaningful result | 183 | 1.68 |
| Total | 10876 | 100.00 |
| **Fine Motor Development** | **Freq** | **Percent** |
| New concern | 54 | 0.50 |
| Previous concern | 68 | 0.63 |
| No concern | 10542 | 96.93 |
| No meaningful result | 212 | 1.95 |
| Total | 10876 | 100.00 |
| **Vision Development** | **Freq** | **Percent** |
| New concern | 76 | 0.70 |
| Previous concern | 163 | 1.50 |
| No concern | 9814 | 90.24 |
| No meaningful result | 823 | 7.57 |
| Total | 10876 | 100.00 |
| **Hearing Development** | **Freq** | **Percent** |
| New concern | 99 | 0.91 |
| Previous concern | 105 | 0.97 |
| No concern | 9827 | 90.35 |
| No meaningful result | 845 | 7.77 |
| Total | 10876 | 100.00 |
